# Adverse pregnancy outcomes and coronary artery disease risk: A negative control Mendelian randomization study

**DOI:** 10.1101/2024.05.11.24307192

**Authors:** Tormod Rogne, Dipender Gill

## Abstract

**Background:** Adverse pregnancy outcomes are predictive for future cardiovascular disease risk, but it is unclear whether they play a causal role. We conducted a Mendelian randomization study with males as a negative control population to estimate the associations between genetic liability to adverse pregnancy outcomes and risk of coronary artery disease.

**Methods:** We extracted uncorrelated (R^2^<0.01) single-nucleotide polymorphisms strongly associated (p-value<5e-8) with miscarriage, gestational diabetes, hypertensive disorders of pregnancy, preeclampsia, placental abruption, poor fetal growth and preterm birth from relevant genome-wide association studies. Genetic associations with risk of coronary artery disease were extracted separately for females and males. The main analysis was the inverse-variance weighted analysis, while MR Egger, weighted median and weighted mode regression, bidirectional analyses, and the negative control population with males were sensitivity analyses to evaluate bias due to genetic pleiotropy.

**Results:** The number of cases for the adverse pregnancy outcomes ranged from 691 (182,824 controls) for placental abruption to 49,996 (174,109 controls) for miscarriage, and there were 22,997 (310,499 controls) and 54,083 (240,453 controls) cases of coronary artery disease for females and males, respectively. We observed an association between genetic liability to hypertensive disorders of pregnancy and preeclampsia, and to some extent gestational diabetes, poor fetal growth and preterm birth, with an increased risk of coronary artery disease among females, which was supported by the MR Egger, weighted median and weighted mode regressions. However, in the negative control population of males, we observed largely the same associations as for females.

**Conclusions:** The associations between adverse pregnancy outcomes and coronary artery disease risk were likely driven by confounding (e.g., shared genetic liability). Thus, our study does not support the hypothesis that adverse pregnancy outcomes are causal risk factors for cardiovascular diseases.

## INTRODUCTION

Adverse pregnancy outcomes such as preterm birth are strongly associated with future maternal risk of cardiovascular diseases.^1–3^ Given that close to a third of females experience an adverse pregnancy outcome during their lifetime, the American Heart Association, among many other societies, are recommending that a history of adverse pregnancy outcomes are taken into account when assessing future risk of cardiovascular diseases.^1–3^

While the *predictive* properties of adverse pregnancy outcomes on future risk of cardiovascular diseases are clear, it is unknown whether they play a *causal* role in the development of cardiovascular diseases, although some biological mechanisms have been hypothesized.^2^ The American Heart Association’s scientific statement includes language such as “[Adverse pregnancy outcomes] increase a woman’s risk of … [cardiovascular diseases]” which may be interpreted as a causal relationship.^1^ An alternative explanation to the link between adverse pregnancy outcomes and risk of cardiovascular diseases is that they reflect confounding due to a common liability.

One way of addressing the question of shared liability is by applying a negative control study within a Mendelian randomization framework.^4^ While genetic liability to an adverse pregnancy outcome can only materialize in females, males carry the same genetic potential (when restricted to the autosomal chromosomes). Thus, if the same associations are observed among females and males, this indicates that the link between genetic liability to adverse pregnancy outcomes and risk of cardiovascular diseases are not due to causal effects of the adverse pregnancy outcomes, but rather due to shared genetic liabilities.

By conducting Mendelian randomization analyses with a negative control design, the aims of this study were to assess whether the adverse pregnancy outcomes outlined by the American Heart Association as screening targets are associated with future risk of coronary artery disease (the leading cause of death in females), and whether these associations are likely to be causal or due to shared genetic liability.

## METHODS

The associations between genetic liability to adverse pregnancy outcomes with the risk of coronary artery disease were assessed by using single nucleotide polymorphisms (SNPs) as genetic instruments for the exposures of interest; a two-sample Mendelian randomization approach. The SNP-coronary artery disease association was divided by the SNP-adverse pregnancy outcome association to provide the Wald ratio.^5^ Three assumptions must be met for an instrument to be valid: 1) it must be associated with the exposure, 2) it cannot be associated with confounders of the exposure-outcome association, 3) and it cannot have an effect on the outcome through other pathways than the exposure.^5^

This study only considered genetic instruments from the autosomal chromosomes. An association between genetic liability to an adverse pregnancy outcome and risk of coronary artery disease among females can be due to a causal effect of the adverse pregnancy outcome, or due to shared genetic etiology (i.e., violating instrumental variable assumptions 2 or 3).

Among males, only the latter is plausible, thus serving as a negative control population (Supplementary Figure 1).

To limit the risk of confounding due to genetic ancestry, only subjects of European ancestry were considered. We evaluated seven adverse pregnancy outcomes that were highlighted by the American Heart Association to be targets for potential screening^1^: miscarriage, gestational diabetes, hypertensive disorders of pregnancy (HDP), preeclampsia, placental abruption, poor fetal growth, and preterm birth. For each adverse pregnancy outcome, we identified the largest genome-wide association study (GWAS) and extracted SNPs that were strongly associated with that adverse pregnancy outcome (p-value < 5e-8) and that were independent of one-another (R^2^ < 0.01) (Table 1). Genetic associations for coronary artery disease – separately for females and males – were extracted from the largest relevant GWAS.

**Table 1.** Genome-wide association studies used as sources for two-sample Mendelian randomization analyses.

| Trait | Study/<br>Source | Setting | Phenotype definition | Cases/controls | Number<br>of SNPs | Variance<br>explained |
| --- | --- | --- | --- | --- | --- | --- |
| **Exposures** |  |  |  |  |  |  |
| Miscarriage | Laisk<br>2020<sup>6</sup> | 13 cohorts,<br>including UK<br>Biobank. | *Cases:* Sporadic miscarriage, either self-reported or retrieved from electronic health records (ICD-10 codes O02.1 or O03).<br>*Controls:* Female controls without registered sporadic miscarriage. | 49,996/174,109 | 10* | 0.2% |
| Gestational<br>diabetes | Elliott<br>2024<sup>7</sup> | FinnGen | *Cases:* Electronic health records (ICD-9 code 6480A; or ICD-10 code O244).<br>*Controls:* Parous females without gestational diabetes. | 12,332/131,109 | 21 | 1.1% |
| HDP | Tyrmi<br>2023<sup>8</sup> | 4 cohorts, including<br>FinnGen and<br>Estonian Biobank | *Cases:* Preeclampsia or other maternal hypertensive disorders. Electronic health records (ICD-10 codes O10, O11, O13-O16; and equivalent ICD-9 and ICD-8 codes.)<br>*Controls:* Parous females without preeclampsia or maternal hypertensive disorders. | 15,200/115,007 | 10 | 0.4% |
| Preeclampsia | Tyrmi<br>2023<sup>8</sup> | 4 cohorts, including<br>FinnGen and<br>Estonian Biobank | *Cases:* Preeclampsia, eclampsia or preeclampsia superimposed on chronic hypertension. Electronic health records (ICD-10 codes O11, O14.0, O14.1, O14.9, O15.0, O15.1, O15.2; and equivalent ICD-9 and ICD-8 codes.)<br>*Controls:* Parous female without preeclampsia or maternal hypertensive disorders. | 16,743/271,306 | 9 | 0.3% |
| Placental<br>abruption | FinnGen,<br>release<br>10<sup>9</sup> | FinnGen | *Cases:* Electronic health records (ICD-9 code 6412; or ICD-10 code O45). *Controls:* Females without mentioned ICD codes. | 691/182,824 | 11* | 1.4% |
| Poor fetal<br>growth | FinnGen,<br>release<br>10<sup>9</sup> | FinnGen | *Cases:* Electronic health records (ICD-9 code 6565A or 6565B; or ICD-10 code O36.5).<br>*Controls:* Females without mentioned ICD codes. | 4,054/226,256 | 12* | 0.6% |
| Preterm birth | Solé-Navais<br>2022<sup>10</sup> | 18 cohorts,<br>including deCODE<br>and UK Biobank. | Singleton live birth with spontaneous onset. *Cases:* Delivery <259 days or ICD-10 code O60.<br>*Controls:* Delivery between 273 to 294 days. | 15,419/217,871 | 5 | 0.2% |
| **Outcome** |  |  |  |  |  |  |
| Coronary<br>artery<br>disease | Aragam<br>2022<sup>11</sup> | 10 cohorts,<br>including deCODE<br>and UK Biobank | *Cases:* Electronic health records (primarily used ICD-10 codes I20-I25).<br>*Controls:* Either non-coronary artery disease subjects in cohort, or population-based controls. | Females: 22,997/310,499<br>Men: 54,083/240,453<br>Both: 181,522/1,165,690 | 112 (bi-<br>directional<br>analyses) | 1.5% (bi-<br>directional<br>analyses) |

All genome-wide association studies on the adverse pregnancy outcomes were conducted on females only. \*P-value threshold was 5e-6 (instead of 5e-8). HDP, hypertensive disorders of pregnancy; ICD, International Classification of Diseases; SNPs, single nucleotide polymorphisms.

Separately for females and males, we used the inverse-variance weighted (IVW) method to calculate the combined association across the Wald ratios for all SNPs for a given adverse pregnancy outcome-coronary artery disease pair.^5^ This method assumes all genetic instruments to be valid. Additionally, we conducted three sensitivity analyses that allow for some invalid instruments; weighted mode, weighted median and MR Egger regression.^5^

Finally, we conducted bidirectional analyses, i.e., evaluating the association between coronary artery disease and risk of the adverse pregnancy outcomes. Given that the clinical onset of coronary artery disease generally occurs later in life compared with the adverse pregnancy outcomes, the same direction of association in the bidirectional analyses was interpreted as an indication of shared genetic etiology (i.e., bias due to pleiotropy). For the bidirectional analyses, we used genetic instruments and genetic associations from the genome-wide association analysis that included both sexes.

The TwoSampleMR (version 0.5.7) package in R (version 4.2.0) was used for all analyses. Explained variance was estimated using the *get_r_from_bsen()* function in the TwoSampleMR package, with effective simple sizes calculated using the *effective_n()* function. Estimates were multiplied with 0.693 (= log_e_ 2) to present the results as the odds ratio of coronary artery disease per doubling in the prevalence of the genetically predicted adverse pregnancy outcome under study. Presence of 95% confidence intervals not including the null were interpreted as statistically significant associations (equal to p-value <0.05). We only used publicly available data with relevant ethical approvals.

## RESULTS

The number of cases for the adverse pregnancy outcomes ranged from 691 (182,824 controls) for placental abruption to 49,996 (174,109 controls) for miscarriage, while the explained variance ranged from 0.2% for miscarriage and preterm birth to 1.4% for placental abruption (Table 1). For coronary artery disease, 22,997 (310,499 controls) and 54,083 (240,453 controls) cases were included for females and males, respectively.

Among females, genetic liability to HDP and preeclampsia were significantly associated with an increased risk of coronary artery disease, and there was a tendency of an increased risk of coronary artery disease with a genetic liability to miscarriage, gestational diabetes, poor fetal growth and preterm birth (Figure 1). There was no indication of a link between genetic liability to placental abruption and risk of coronary artery disease. The same patterns of association were observed among males (Figure 1).

**Figure 1.**
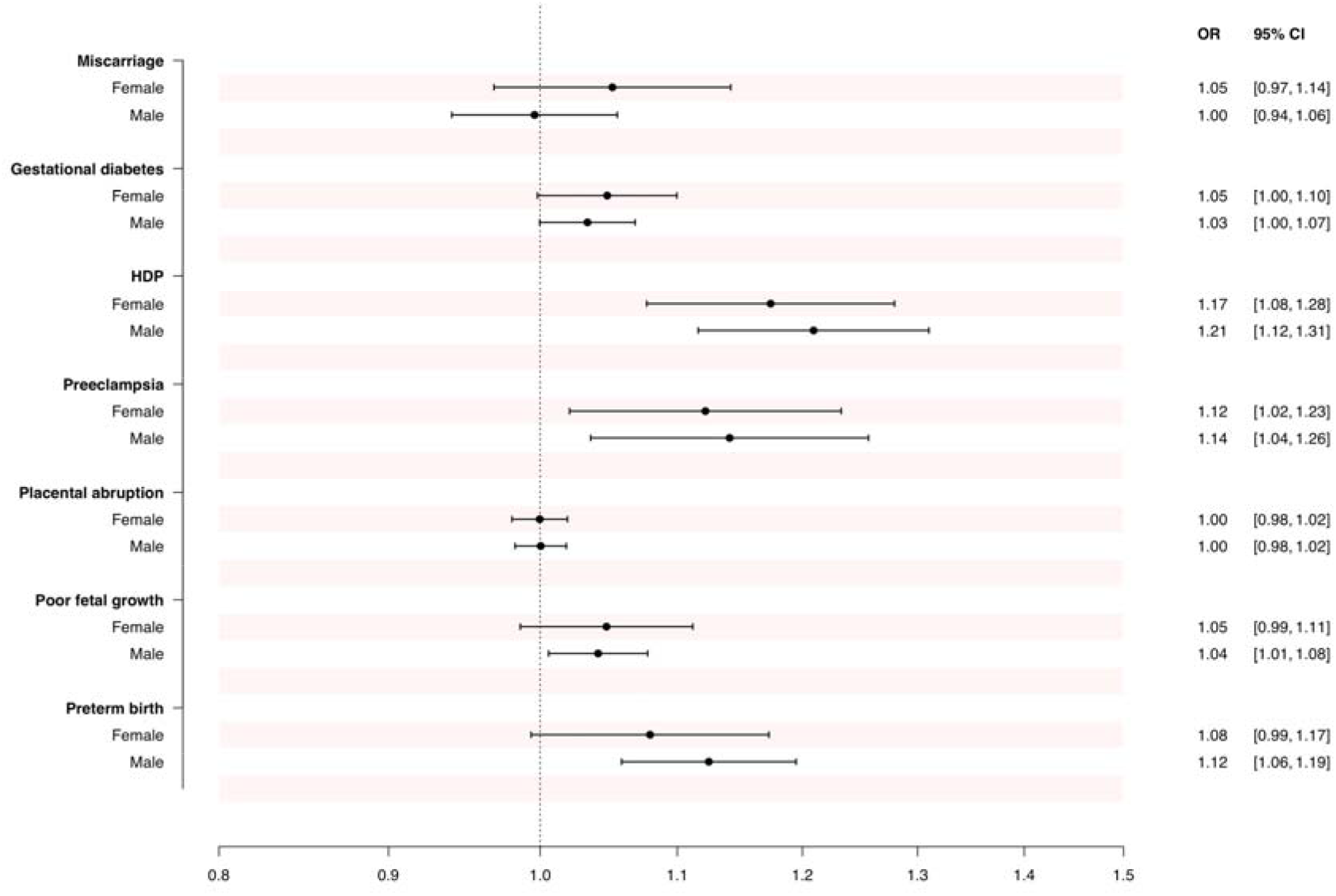
Associations between genetic liability to adverse pregnancy outcomes and the risk of coronary artery disease, among females and males. *Legend:* Results from the inverse-variance weighted analyses, expressed as OR of coronary artery disease per doubling in the prevalence of the genetically predicted adverse pregnancy outcome. CI, confidence interval; HDP, hypertensive disorders of pregnancy (including preeclampsia); OR, odds ratio.

The weighted median, weighted mode and MR Egger regressions among females generally supported the inverse-variance weighted analyses (Supplementary Figure 2). In the bidirectional analyses, genetic liability to coronary artery disease was associated with a significantly increased risk of HDP, preeclampsia and preterm birth, and a tendency of an increased risk of gestational diabetes (Supplementary Figure 3).

## DISCUSSION

In this two-sample Mendelian randomization study using a negative control population, we observed that the patterns of association between genetic liability to adverse pregnancy outcomes and risk of coronary artery disease was similar among females and males, which strongly indicates that these associations were due to shared genetic liability.

The observed positive associations between genetic liability to HDP and preeclampsia – and to some extent gestational diabetes, poor fetal growth and preterm birth – with the risk of coronary artery disease among females support previous population-based observational studies.^1,2^ One of these studies included sibling comparisons which addresses some of the potential bias due to shared genetic liability, with similar findings.^2^

Our findings also support the two previous Mendelian randomization studies that have been conducted on this topic. These evaluated the role of genetic liability to HDP on future risk of cardiovascular diseases, and both found a positive association.^12,13^ One of these studies also considered males in a negative control population, where they observed the same findings as in the female-only analyses.^13^ However, no previous genetic epidemiological study have evaluated the role of the other adverse pregnancy outcomes considered in our study; miscarriage, gestational diabetes, placental abruption, poor fetal growth, and preterm birth.

The quantitative tests often used to address concerns of pleiotropy that were included in this study – weighted median, weighted mode, and MR Egger regression – did not indicate presence of pleiotropy. This is in contrast with the bidirectional tests which largely supported the negative control analyses.

This study has several strengths, but also some weaknesses. This is the first Mendelian randomization study to evaluate the associations between genetic liability to miscarriage, gestational diabetes, placental abruption, poor fetal growth, and preterm birth, with the risk of cardiovascular diseases. It is also one of relatively few Mendelian randomization studies applying a negative control design, which was key to establish that the patterns of association were due to shared genetic liability. That said, there may be other biological differences in coronary artery disease risk between the sexes than those related to pregnancy outcomes that could affect our findings. However, the genetic liability to coronary artery disease risk was reported to be very similar for females and males.^11^ Low explained variance of the genetic instruments yielded wide confidence intervals. Another important limitation is that due to data availability we only considered subjects of European ancestry. While this greatly reduces risk of confounding due to population characteristics, it importantly limits generalizability to other population groups.

## Conclusion

Concordant with previous studies, our findings support that HDP and preeclampsia, and to some extent gestational diabetes, poor fetal growth and preterm birth, are important predictors of future risk of coronary artery disease. However, these associations were likely confounded by shared genetic liability, and our study does not support the hypothesis that adverse pregnancy outcomes are causal risk factors for cardiovascular diseases.

## Data Availability

All data used in the present study are publicly available.

## REFERENCES

1 Parikh NI, Gonzalez JM, Anderson CAM, Judd SE, Rexrode KM, Hlatky MA, et al. Adverse Pregnancy Outcomes and Cardiovascular Disease Risk: Unique Opportunities for Cardiovascular Disease Prevention in Women: A Scientific Statement From the American Heart Association. Circulation 2021;143:. 10.1161/CIR.0000000000000961.

2 Crump C, Sundquist J, McLaughlin MA, Dolan SM, Govindarajulu U, Sieh W, et al. Adverse pregnancy outcomes and long term risk of ischemic heart disease in mothers: national cohort and co-sibling study. BMJ 2023:e072112. 10.1136/bmj-2022-072112.

3 Lewey J, Beckie TM, Brown HL, Brown SD, Garovic VD, Khan SS, et al. Opportunities in the Postpartum Period to Reduce Cardiovascular Disease Risk after Adverse Pregnancy Outcomes: A Scientific Statement from the American Heart Association. Circulation 2024;149:E330–46. 10.1161/CIR.0000000000001212.

4 Skrivankova VW, Richmond RC, Woolf BAR, Davies NM, Swanson SA, VanderWeele TJ, et al. Strengthening the reporting of observational studies in epidemiology using mendelian randomisation (STROBE-MR): explanation and elaboration. BMJ 2021;2:n2233. 10.1136/bmj.n2233.

5 Sanderson E, Glymour MM, Holmes M V., Kang H, Morrison J, Munafò MR, et al. Mendelian randomization. Nat Rev Methods Prim 2022;2:6. 10.1038/s43586-021-00092-5.

6 Laisk T, Soares ALG, Ferreira T, Painter JN, Censin JC, Laber S, et al. The genetic architecture of sporadic and multiple consecutive miscarriage. Nat Commun 2020;11:5980. 10.1038/s41467-020-19742-5.

7 Elliott A, Walters RK, Pirinen M, Kurki M, Junna N, Goldstein JI, et al. Distinct and shared genetic architectures of gestational diabetes mellitus and type 2 diabetes. Nat Genet 2024. 10.1038/s41588-023-01607-4.

8 Tyrmi JS, Kaartokallio T, Lokki AI, Jääskeläinen T, Kortelainen E, Ruotsalainen S, et al. Genetic Risk Factors Associated With Preeclampsia and Hypertensive Disorders of Pregnancy. JAMA Cardiol 2023;8:674. 10.1001/jamacardio.2023.1312.

9 Kurki MI, Karjalainen J, Palta P, Sipilä TP, Kristiansson K, Donner KM, et al. FinnGen provides genetic insights from a well-phenotyped isolated population. Nature 2023;613:508–18. 10.1038/s41586-022-05473-8.

10 Solé-Navais P, Flatley C, Steinthorsdottir V, Vaudel M, Juodakis J, Chen J, et al. Genetic effects on the timing of parturition and links to fetal birth weight. Nat Genet 2023;55:559–67. 10.1038/s41588-023-01343-9.

11 Aragam KG, Jiang T, Goel A, Kanoni S, Wolford BN, Atri DS, et al. Discovery and systematic characterization of risk variants and genes for coronary artery disease in over a million participants. Nat Genet 2022;54:1803–15. 10.1038/s41588-022-01233-6.

12 Rayes B, Ardissino M, Slob EAW, Patel KHK, Girling J, Ng FS. Association of Hypertensive Disorders of Pregnancy with Future Cardiovascular Disease. JAMA Netw Open 2023;6:E230034. 10.1001/jamanetworkopen.2023.0034.

13 Tschiderer L, van der Schouw YT, Burgess S, Bloemenkamp KWM, Seekircher L, Willeit P, et al. Hypertensive disorders of pregnancy and cardiovascular disease risk: a Mendelian randomisation study. Heart 2023:heartjnl-2023-323490. 10.1136/heartjnl-2023-323490.

