## Supplementary for "Adverse pregnancy outcomes and coronary artery disease risk: A negative control Mendelian randomization study"

**SUPPLEMENTARY FIGURES**

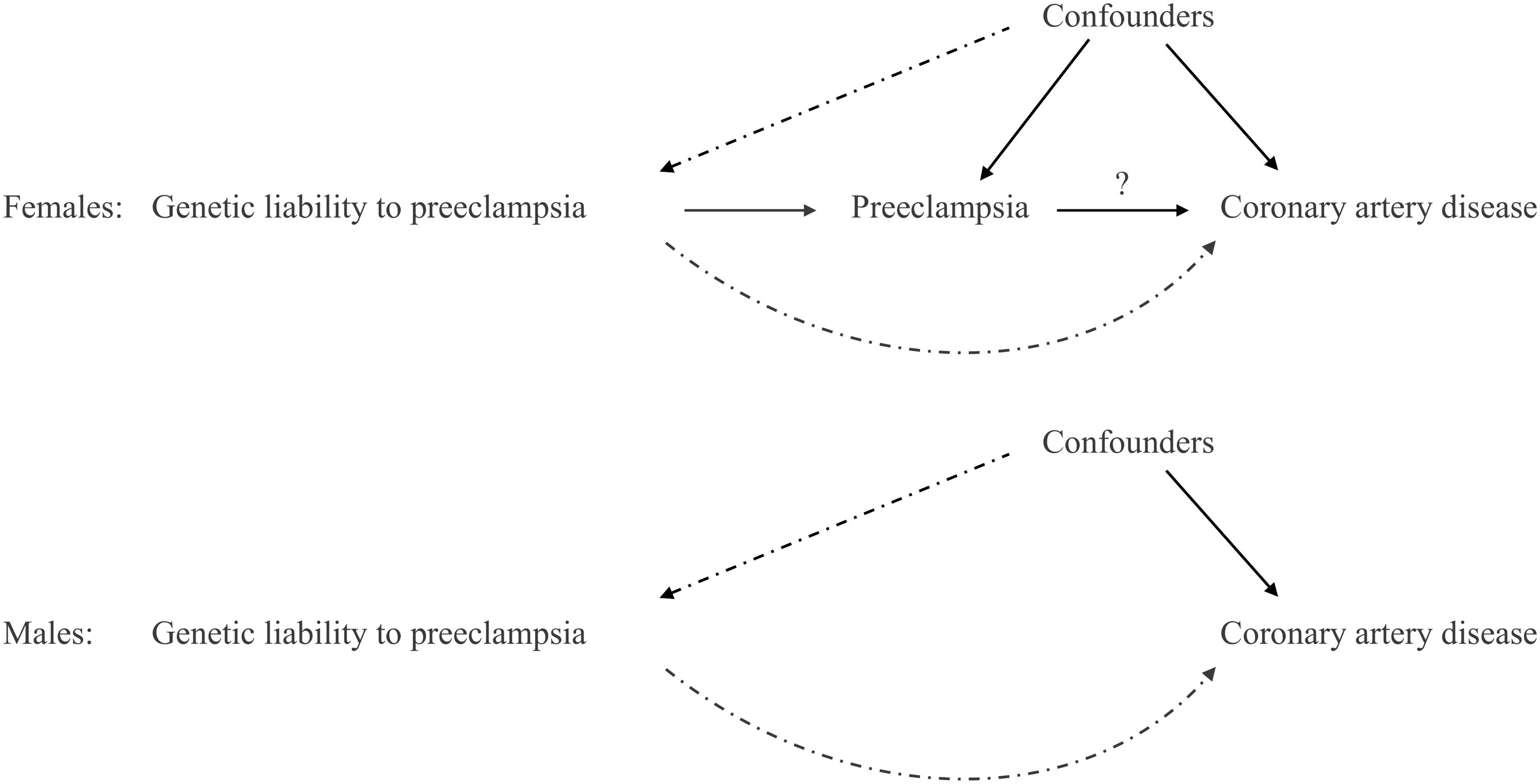

**Supplementary Figure 1.** Directed acyclic graph of adverse pregnancy outcomes and risk of coronary artery disease.

*Legend:* Solid arrows represent suspected causal effects under the hypothesis that preeclampsia may have a causal effect on coronary artery disease, dashed arrows represent potential biasing pathways, and arrow with question mark represent the association under study. Preeclampsia is used as an example of an adverse pregnancy outcome. The instrumental variable assumptions would be violated if genetic liability to preeclampsia affects coronary artery disease other than through preeclampsia, and if it is associated with any confounders. In the negative control population of men, where preeclampsia cannot occur, any association between genetic liability to preeclampsia and risk of coronary artery disease is indicative of violation of the instrumental variable assumptions.

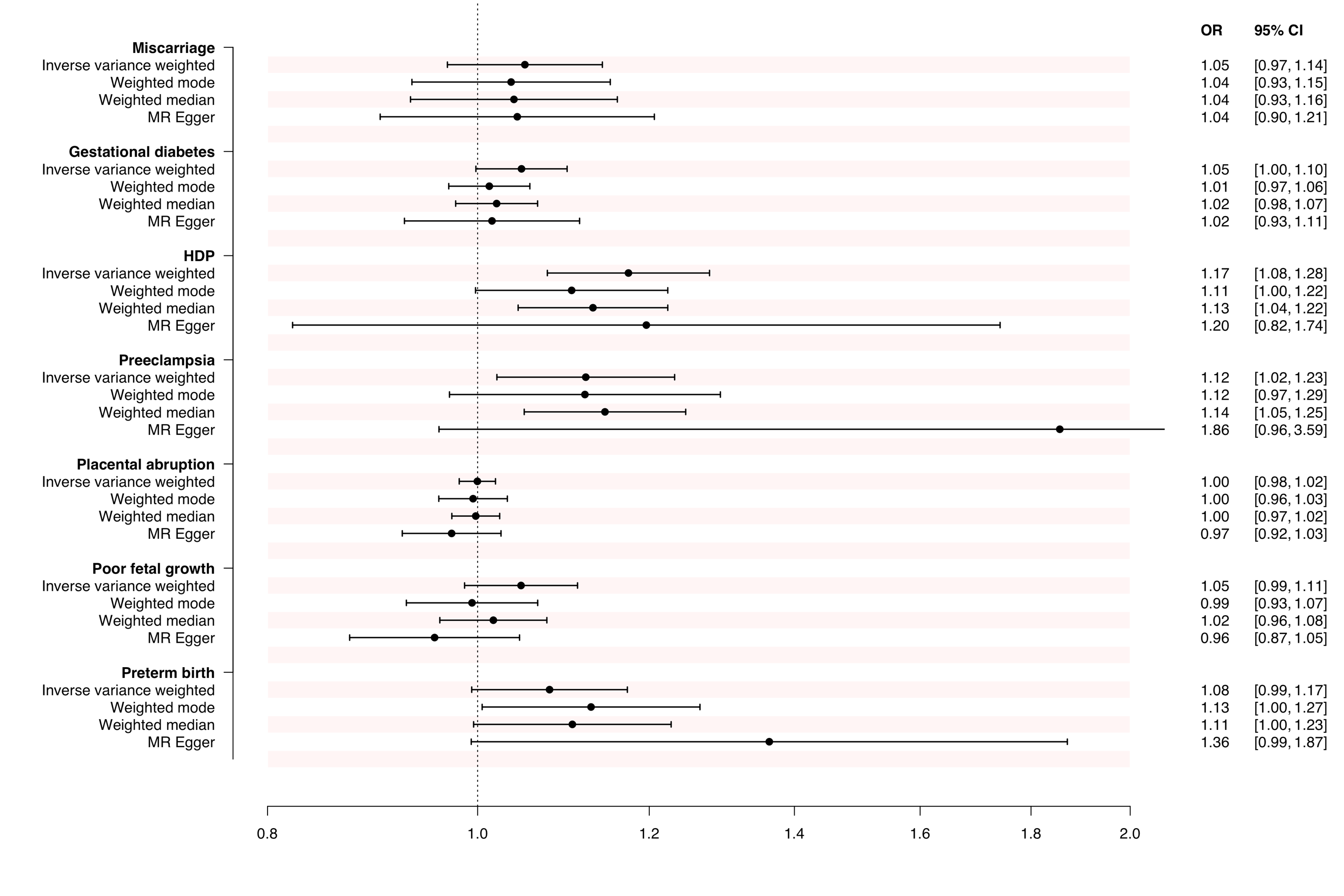

**Supplementary Figure 2.** Statistical sensitivity analyses for the Mendelian randomization associations between genetic liability to adverse pregnancy outcomes and the risk of coronary artery disease among females.

*Legend:* Results expressed as OR of coronary artery disease per doubling in the prevalence of the genetically predicted adverse pregnancy outcome. CI, confidence interval; HDP, hypertensive disorders of pregnancy (including preeclampsia); OR, odds ratio.

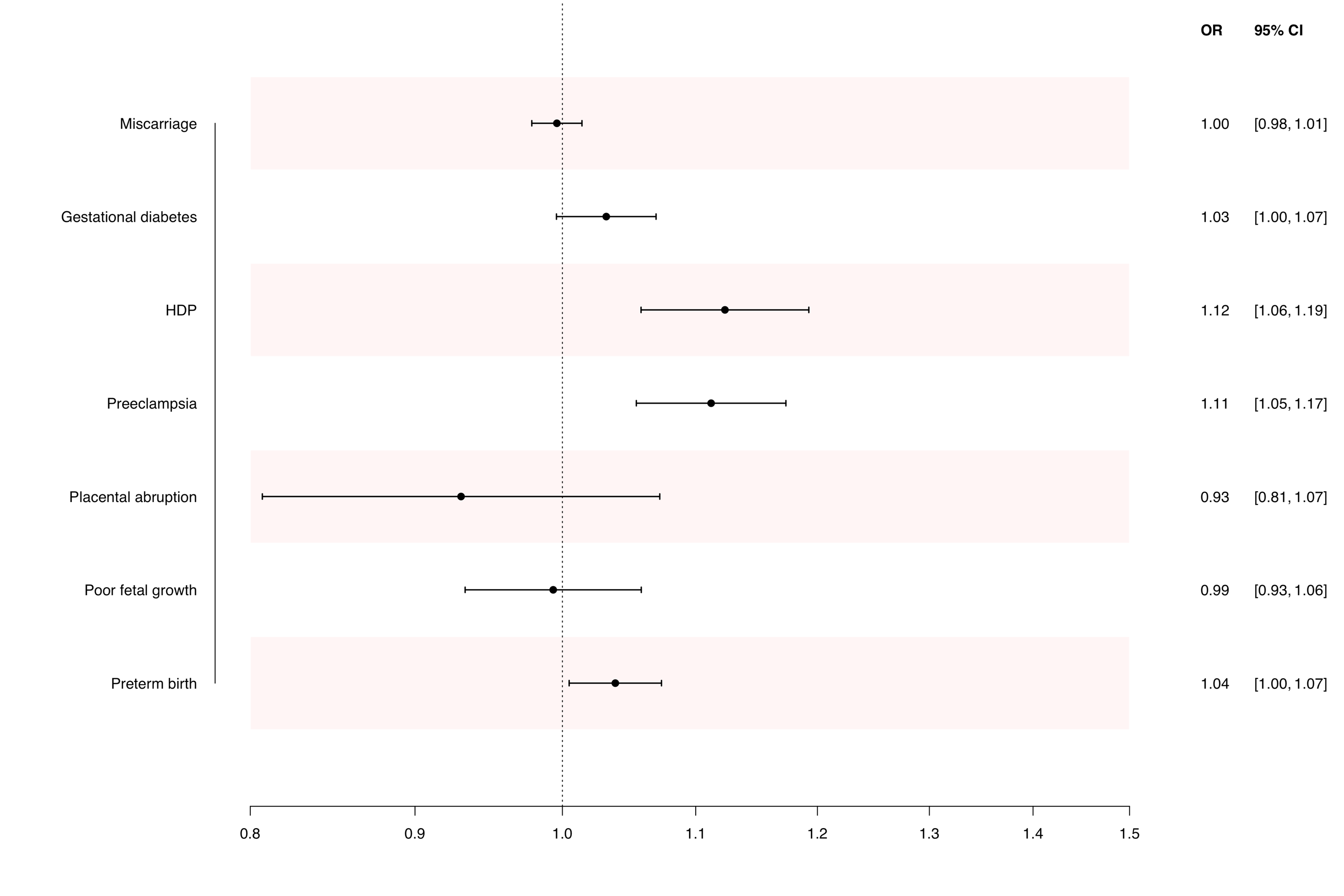

**Supplementary Figure 3.** Bidirectional sensitivity analyses of the Mendelian randomization associations between genetic liability to coronary artery disease and the risk of adverse pregnancy outcomes among females.

*Legend:* Results expressed as OR of the adverse pregnancy outcome per doubling in the prevalence of genetically predicted coronary artery disease. CI, confidence interval; HDP, hypertensive disorders of pregnancy (including preeclampsia); OR, odds ratio.
